# *In vitro* and *in situ* analysis of a novel copper-based antimicrobial surface coating designed to reduce the microbial bioburden of high touch surfaces in a hospital environment

**DOI:** 10.1101/2025.06.30.25330568

**Authors:** Gavin Ackers-Johnson, Joseph M Lewis, Patricia Killen, Victor Bellido-Gonzalez, Lara Maroto-Diaz, Dermot Monaghan, Jason Eite, Rick Spencer, Ameen Yunus, Danielle McLaughlan, Amy Doyle, Tim Neal, Stacy Todd, Adam P. Roberts

## Abstract

The hospital environment plays a crucial role in healthcare associated infections. Current cleaning protocols to address this are costly and labour intensive, with further complications related to compliance, efficiency and the environmental impact of cleaning agents.

We developed a nano-structured coating, iC-nano^™^, which exhibits high antimicrobial activity. The 73 unique formulations assessed showed a range of activity *in vitro*. The best coatings displayed high activity within 15 minutes and were equally effective against all ESKAPE(e) pathogens except *Enterococcus faecium*, which required an increased contact time.

Analysis within an active hospital environment identified 44% fewer CFUs recovered from coated surfaces, though with some uncertainty in estimates of effect size (1.77-fold [95% CrI: 0.98, 3.13]). The highest counts were observed amongst toilet exits, whilst samples collected on Wednesday had 45% less CFUs than on Monday (0.55-fold [95% CrI: 0.32, 0.91]). Whilst the durability and antimicrobial activity of the coating remained stable, a key concern raised was the aesthetic of the product that gave the appearance of being soiled after sustained use, potentially leading to behaviour change.

Through this study we highlighted the promising potential of antimicrobial surfaces coatings. We suggest that future interventions, be it enhanced cleaning protocols or specialised coatings, should initially focus on locations such as toilet door handles, where the largest recoverable microbial bioburden was observed. Conversely, patient check in screens saw minimal human interaction and are therefore considered a lower priority.

## Introduction

Healthcare associated infections (HAIs) present a substantial challenge to healthcare systems, posing risks to patients, visitors, and healthcare professionals alike. They not only result in increased morbidity and mortality, but also impose financial burdens on healthcare providers (1). The World Health Organization estimates that 7% of patients in high-income and 15% of patients in low- and middle-income countries contract at least one HAI during their hospitalisation (2). Antimicrobial resistance (AMR) further compounds this issue. Being the direct cause of 1.27 million, and contributing towards 4.95 million deaths globally in 2019, AMR is quickly becoming one of the top threats to global public health (3). Indeed, the majority of HAIs are attributed to the ESKAPE(e) pathogens (*Enterococcus faecium, Staphylococcus aureus, Klebsiella pneumoniae, Acinetobacter baumannii, Pseudomonas aeruginosa, Enterobacter spp*. and *Escherichia coli*) which are frequently associated with multidrug resistance (MDR) and pose the highest risk of mortality (4).

The hospital environment plays a pivotal role in HAI development, with opportunistic bacteria readily identifiable on inanimate surfaces (5). Simply admitting a new patient to a room previously occupied by an infected or colonised individual poses a significant risk for further transmission (6). Similar issues arise in relation to antimicrobial resistance. Due to treatment and cleaning regimens within healthcare settings, there is a prolonged abundance of antimicrobials and cleaning disinfectants. This creates a strong selective pressure for the generation of resistance (7). Just as environmental surfaces act as a reservoir for potentially harmful bacteria, bacteria with relatively low clinical significance have the potential to act as a reservoir of AMR genes, which can spread through the local microbiome via the movement of mobile genetic elements (8). If acquired by a potential pathogen, the treatment of future infections would become increasingly difficult, highlighting the need for effective infection prevention and control (IPC) strategies throughout healthcare infrastructure (9).

Interventions aimed at reducing HAIs, patient colonisation and environmental bioburden, such as cleaning methods involving chemical, mechanical, and human factors, often yield favourable outcomes (10). However, these come with their own challenges. Traditional methods of cleaning involve a range of chemical disinfectants and detergents, including quaternary ammonium compounds, hydrogen peroxide and chlorine-based solutions (11). These microbicidal compounds are not only variably toxic to humans, leading to adverse reactions and occupational illnesses, but have a mounting environmental impact with their presence being detected in surface and ground waters (12). Whilst there is an ever increasing choice of products to aid in decontaminating surfaces, including automated devices, impregnated wipes and novel disinfectants, these often come at an economic cost unsustainable for many settings (13).

Maintaining a completely sterile environment is largely unachievable; even after exposure to powerful microbicides, subsequent recolonisation events occur to fill the empty ecological niche (14). As such, self-sanitising surfaces are of growing interest through their persistent antimicrobial activity. These coated surfaces are typically comprised of metal alloys. Although a range of alloys display potent antimicrobial activity, the nature of the target surface is an important consideration. For example, silver requires moisture for its action (15), whilst titanium dioxide requires exposure to ultraviolet light (16). Copper, meanwhile, maintains its activity in the absence of light and in both aquatic systems and on dry surfaces, making it ideal for use in healthcare settings (17, 18). Furthermore, copper has confirmed antiviral activity and is the only solid antimicrobial material registered as a self-sanitiser by Health Canada and the U.S. Environmental Protection Agency (19).

The aim of this study was to investigate the antimicrobial efficacy of door handles/panels and transparent touch screen covers coated with novel copper-based compounds through *in vitro* laboratory and *in situ* hospital analysis. Compounds were developed with the aim of combining broad-spectrum antimicrobial activity with enhanced mechanical durability and coated onto surfaces utilising magnetron sputtering.

## Methods

### Novel copper-based surface coatings were developed at Gencoa Ltd, Liverpool (20)

*In vitro* antimicrobial analysis of the coatings was facilitated at the Liverpool School of Tropical Medicine. Bacteria (*Enterococcus faecium* NCTC 7171, *Staphylococcus aureus* NCTC 8532, *Klebsiella pneumoniae* NCTC 9633, *Acinetobacter baumannii* NCTC 12156, *Pseudomonas aeruginosa* NCTC 13437, *Enterobacter cloacae* NCTC 10005 *and Escherichia coli* NCTC 86) were cultured in 10ml LB broth overnight at 37°C in a shaking incubator. The optical density at 600nm wavelength (OD600) of the overnight culture was measured, whilst 1ml of overnight culture was centrifuged at 14,000 rpm for ten minutes. The supernatant was discarded, and the pellet resuspended in phosphate buffered saline (PBS) to an OD600 of 0.5. 2cm^2^ coupons of coated material were cleaned with 70% ethanol and left to air dry. 100ul of resuspended bacterial culture was added to each coupon in five 20ul drops. Surfaces were incubated at room temperature for a variety of contact times before reaching the final benchmark of one hour. Given the primary sites of installation would be door handles, which could be interacted with multiple times in short succession, a contact time of 24 hours was deemed insufficient. To recover bacteria each coupon was added to an individual 50ml falcon tube containing 5ml maximum recovery diluent. Tubes were vortexed at maximum speed for 30 seconds. Recovered solutions were serially diluted up to 10^−4^ and plated onto tryptic soy agar (TSA). TSA plates were incubated overnight at 37°C. Colony forming units (CFUs) were counted to assess the quantity of bacteria recovered.

*In situ* analysis of coated and uncoated door handles/panels was facilitated on the same infectious diseases ward at the Liverpool Royal Hospital, which contained 19 en-suite bedrooms and six utility/staff/other rooms included in the study. Samples were collected on Mondays and Wednesdays, with 899 uncoated samples initially collected between 10^th^ July 2023 and 6^th^ November 2023. The first batch of 14 coated surfaces (5 push plates and 9 handles) were installed 5^th^ December 2023 and subsequently removed 10^th^ January 2024. They were positioned in two utility rooms, the main ward entrance, the public toilet and three bedrooms. The second batch of 29 coated surfaces (7 push plates and 22 handles) were installed 5^th^ February 2024 and were subsequently removed 28^th^ February 2024. They were positioned in the same locations as the previous 14 coated surfaces, in addition to five extra bedrooms. 158 coated samples were collected in total.

Coated and uncoated transparent screens were assessed utilising outpatient check-in screens. Six screens were included, two of which were coated whilst four were left uncoated. Swab samples were collected on Wednesdays at four time points: 09:00, 11:00, 13:00 and 15:00. Between 8^th^ August 2023 and 6^th^ December 2023, 330 uncoated and 132 coated samples were processed.

Sample collection utilised 3D printed 25cm^2^ templates and cotton swabs premoistened in 5ml neutralising buffer. The target site was swabbed in four directions across the template to ensure complete coverage; top to bottom, left to right, top-left to bottom-right and top-right to bottom-left. Samples were transported to the Liverpool School of Tropical Medicine, where the swabs and neutralising buffer were processed in a Stomacher 80 Biomaster (Seward, Worthing, United Kingdom) at maximum speed for two minutes, with 500µl of diluent plated onto 5% sheep’s blood agar, followed by a subsequent 48-hour incubation at 37°C. Colonies were counted and colony forming units/cm^2^ calculated.

We used the brms package (21) in R (v4.5.0) to fit two separate Bayesian zero-inflated negative binomial regression models to evaluate:

1. the fixed effects of the day of sampling (Monday vs Wednesday) and surface type (coated vs uncoated), with random intercepts included for door location and sampling date. Door location was included to account for variation between physical sites with variable usage. Sampling date was included to account for potential correlations among samples collected on the same day (e.g. staff cleaning schedules or transient seasonal variations impacting the ward environment).
2. the fixed effects of touchscreen sampling time as a categorical variable (09:00, 11:00, 13:00 and 15:00; 09:00 used as the reference level) and surface type (coated vs uncoated).

The raw CFU data was modelled using a log link function for the expected count, and an identity link for the zero-inflation probability. Convergence of MCMC chains in brms was assessed by inspection of trace plots and the Gelman-Rubin statistic 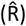 being close to 1. Parameter values are expressed as posterior mean with 95% credible intervals (CrI). Model outputs are available in Supplementary Table A1.

## Results

### *In vitro* analysis of coatings

73 unique formulations of coated material were assessed (Figure 1), which showed a range of activity from complete kill through to no effect on the growth of *S. aureus* within 1 hour. Assessment of iCn-010 and iCn-013 identified large CFU reductions (99.4%) in as little as 15 minutes, with further tests against all ESKAPE(e) pathogens showing similar results (Supplementary Figure A1). The sole exception was *E. faecium*, where both a clinical and reference strain NCTC 7171 required a longer contact time of up to 4 hours for comparable reductions.

**Figure 1.**
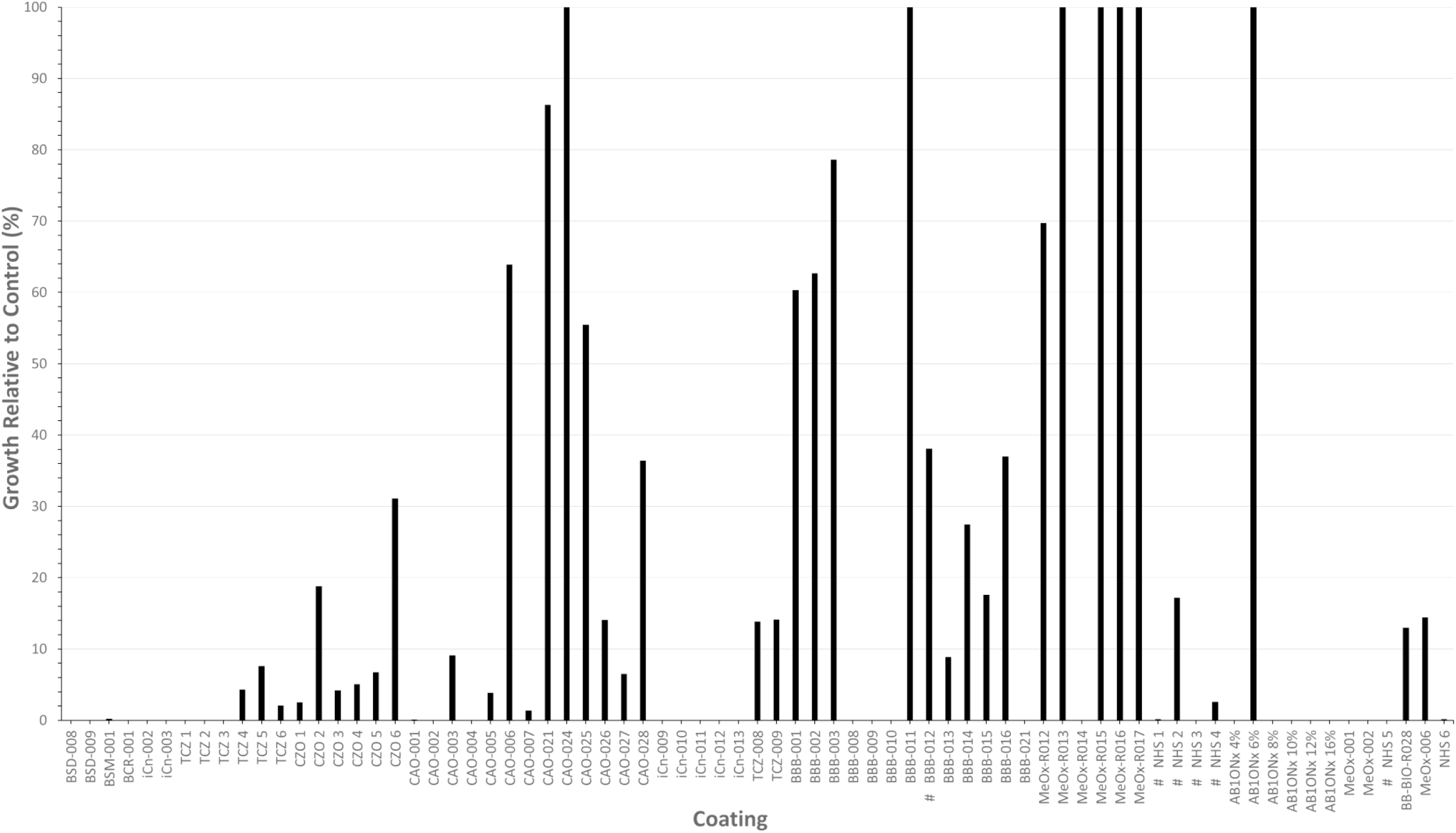
The percentage of *Staphylococcus aureus* colony forming units recovered from coatings relative to the control after a 1-hour incubation at room temperature. Coatings later tested *in situ* at the hospital are marked with a (#). BBB-012 was used to coat the outpatient check in screens, whilst NHS 1-5 were used for handles and door panels on the ward.

A common phenomenon identified in preliminary testing was the appearance of fingerprint ‘grease marks’ on door panels, the visual outcome of the reaction between copper and organic material. Anecdotally, this led to people actively avoiding touching the seemingly heavily soiled surfaces. This was observed extensively where alcohol hand gel was in use, accelerating the visual degradation of the coated surface. *In vitro* analysis revealed that whilst overnight incubation in alcohol hand gel reduced the efficacy of the coating to a minor extent, the primary concern was the distinctive impact it had on visual acceptability. Given the prevalence of alcohol hand gel in hospital settings, extensive work was facilitated to minimise this visual effect. Whilst the coatings with a high copper load had the highest antimicrobial efficacy, they were the most susceptible to visual degradation.

### *In situ* analysis of coated door handles and panels

Median values on all collection dates except four (three uncoated and one coated) were less than 10 CFU/cm^2^. However, a sizeable number of individual points were much higher (Figure 2). Model one estimated a posterior mean of 0.57 on the log scale for the effect of uncoated handles (95% CrI: - 0.02, 1.16), a 77% increase in CFUs compared to coated handles (1.77-fold [95% CrI: 0.98, 3.19]). The posterior distribution indicated a 97% probability that uncoated handles have higher bacterial loads, providing strong evidence of a positive effect of coated surfaces. Random intercepts for sampling date (SD = 0.61 [95% CrI: 0.41, 0.87]) and door location (SD = 1.25 [95% CrI: 0.63, 2.54]) captured variation across sampling times and locations.

**Figure 2.**
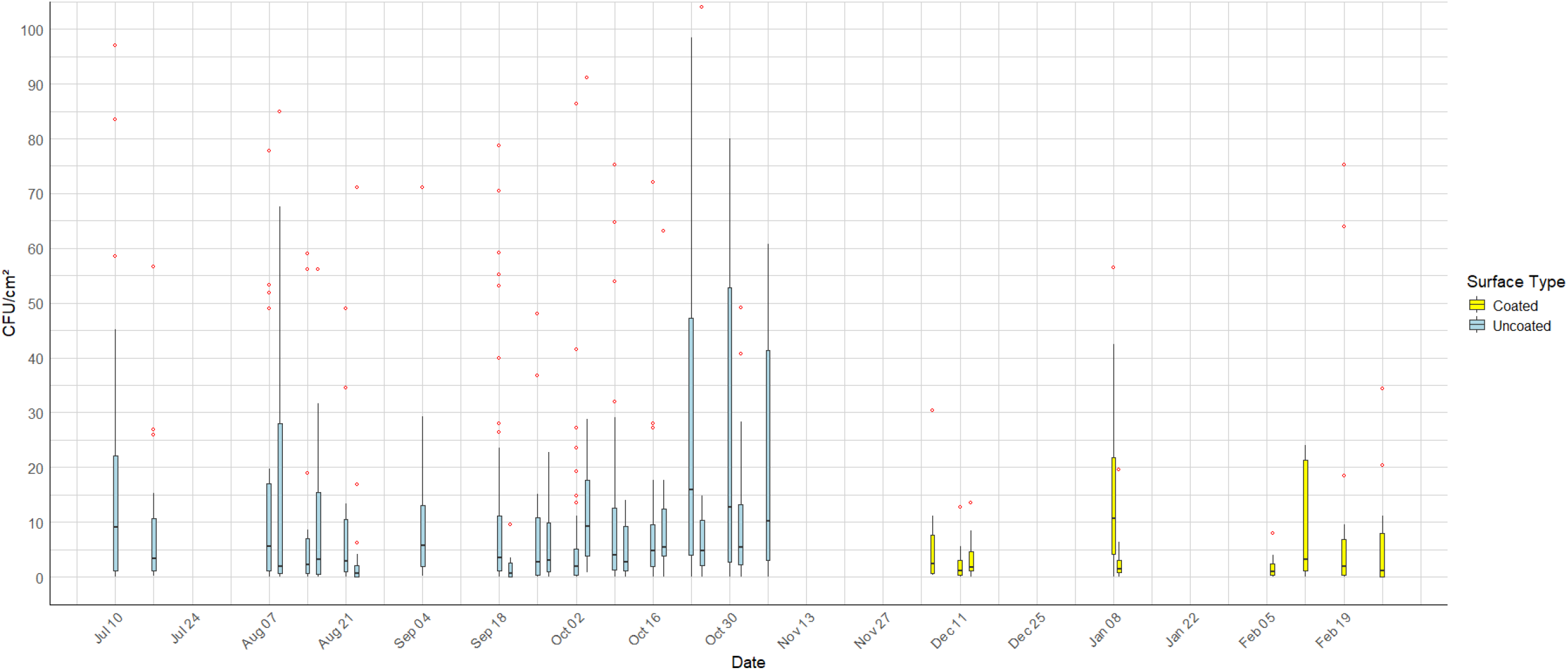
The CFU/cm^2^ recovered at each time point from door handles and panels on the Infectious Diseases ward at the Liverpool Royal Hospital. The boxplot displays the median (horizontal line), the interquartile range (box spans the 25th to 75th percentiles), and whiskers that extend to the smallest and largest values within 1.5 times the IQR from the lower and upper quartiles, respectively. Observations outside this range are shown as individual outlier points, with points above 100 CFU/cm^2^ hidden from view.

Sample sites were primarily located in single occupancy, en-suite, inpatient bedrooms and their respective adjoining rooms. Median CFU/cm^2^ values showed toilets frequently had the highest levels of recoverable bacteria, with high variability observed (Figure 3a). The 95% CrIs for the random effect estimates of door locations suggested that toilet doors were likely to have CFU counts higher than the average across all sites. However, overlapping CrIs with other locations in the trace plot indicated uncertainty about whether this difference was statistically meaningful (Figure 3b).

**Figure 3.**
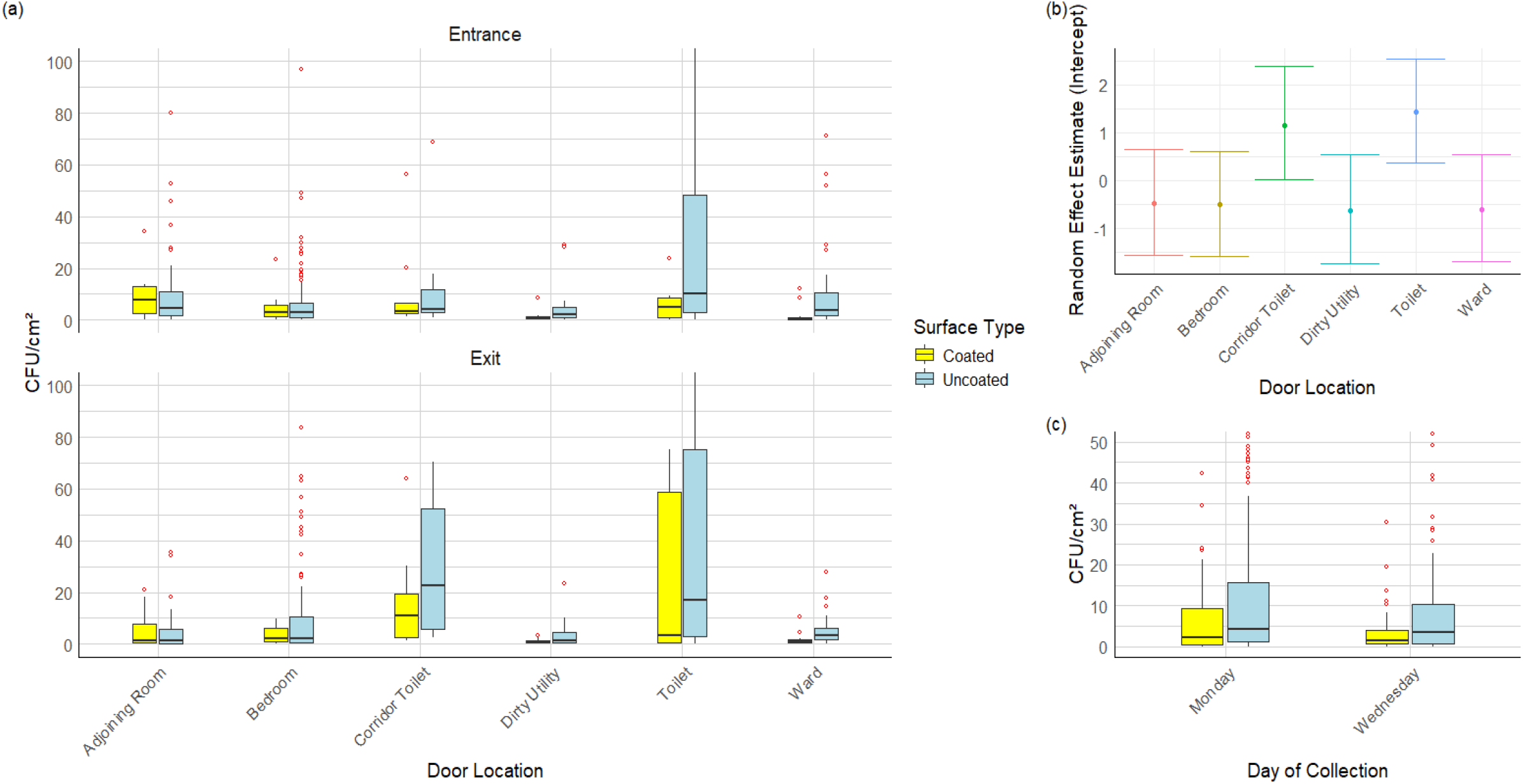
The CFU/cm^2^ recovered from (a) the entrance and exit of each sample location; and (c) from each weekday of sample collection. The locations sampled were Adjoining Rooms (situated between two bedrooms), Bedrooms, Toilets (en-suite), Corridor Toilets, Dirty Utility Room and the main entrance to the Ward. The boxplots display the median (horizontal line), the interquartile range (box spans the 25th to 75th percentiles), and whiskers that extend to the smallest and largest values within 1.5 times the IQR from the lower and upper quartiles, respectively. Observations outside this range are shown as individual outlier points, with points above (a) 100 CFU/cm^2^ and (c) 50 CFU/cm^2^ hidden from view. (b) represents the random effect estimate for each door location, with error bars indicating 95% credible intervals.

Most samples were collected on Monday mornings. However, a selection were collected on Wednesdays to identify any variation across different days of the week (Figure 3c). The posterior mean for the effect of sampling on Wednesday compared to Monday was -0.59 on the log scale (95% CrI: -1.13, -0.09), a 45% decrease in CFUs (0.55-fold [95% CrI: 0.32, 0.91]). The posterior distribution indicated a 99% probability that surfaces on Monday have higher bacterial loads than on Wednesday, suggesting a strong mid-week reduction in surface contamination.

### *In situ* analysis of coated surfaces installed on hospital outpatient check in screens

CFUs recovered from check in screens were lower than those from door handles/panels, with median values at 11:00, 13:00 and 15:00 all close to 1 CFU/cm^2^ for both coated and uncoated surfaces (Figure 4). More colonies were recovered around the 09:00 sample point, with a noticeable median reduction on the coated screens. Model two highlighted this trend, with bacterial load substantially reduced by 81% at 13:00 (0.19-fold [95% CrI: 0.11, 0.31]) and 51% at 15:00 (0.49-fold [95% CrI: 0.29, 0.79]) relative to 09:00. The estimated 30% reduction at 11:00 (0.70-fold [95% CrI: 0.43, 1.14]) was less certain. Whilst the model estimated uncoated screens had 28% higher CFU counts, the effect of the surface coating was uncertain (1.28-fold [95% CrI: 0.86, 1.89]).

**Figure 4.**
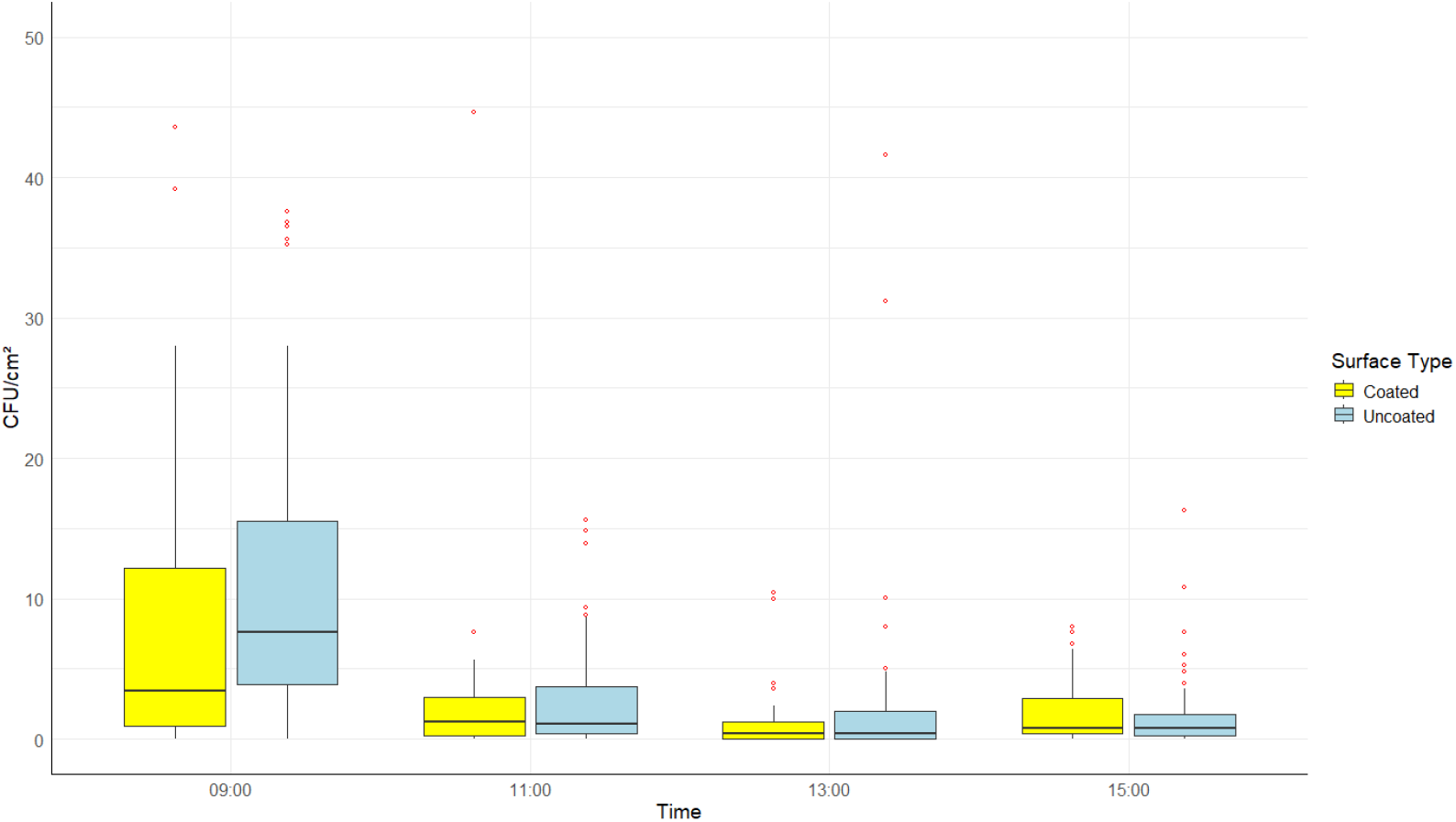
The CFU/cm^2^ recovered from coated and uncoated outpatient check in screens at the Liverpool Royal Hospital. Samples were collected on Wednesdays at 09:00, 11:00, 13:00 and 15:00. The boxplot displays the median (horizontal line), the interquartile range (box spans the 25th to 75th percentiles), and whiskers that extend to the smallest and largest values within 1.5 times the IQR from the lower and upper quartiles, respectively. Observations outside this range are shown as individual outlier points, with points above 30 CFU/cm^2^ hidden from view.

## Discussion

### *In vitro* analysis of coatings

Standard methodology for assessing the antimicrobial efficacy of plastics and other non-porous surfaces utilises the ISO 22196:2011 standard. Despite being the gold standard, it has several pitfalls (22). As such, protocol modifications are frequently observed (23). Most critically, it has been demonstrated to poorly reflect real-life scenarios, utilising a high temperature, humidity and contact time of 24 hours (24). We demonstrated a wide scope of antimicrobial activity using modified protocols designed to mimic *in situ* conditions. The efficacy of different coating compositions varied significantly, from having no effect to complete kill of the bacteria within one hour. As was expected with copper being a well-known antimicrobial agent (25), compositions with a high copper load were generally more successful. However, we aimed to produce coatings which combined both antimicrobial activity and mechanical durability for use in practical applications. Our ‘NHS’ coatings successfully fulfilled these parameters when testing *in vitro* and were deemed suitable for *in situ* assessment. Further constraining our coated surfaces was their aesthetic. Installing the surfaces in a public-facing, new hospital, their visual appearance played an important role. It was essential that the surfaces remained not only clinically clean, but also visually clean as to not adversely affect peoples’ behaviour and hence the surfaces’ functionality.

ISO 22196:2011 guidelines utilise *Staphylococcus aureus* or *Escherichia coli* as a reference for gram-positive and gram-negative organisms respectively (22). For our high throughput tests we chose to focus on *Staphylococcus aureus*, with our previous study at the same location identifying *Staphylococcus* spp. as one of the primary colonisers of door handles (5). For our high-performance coatings, we assessed and observed high efficacy against all ESKAPE(e) pathogens except *E. faecium*. Both a MDR clinical strain and NCTC 7171 reference strain required a contact time of up to four hours (increased from one hour) to achieve comparable CFU reductions. *E. faecium* is known to exhibit intrinsic copper tolerance, with homeostasis maintained through the *cop* operon (26). Furthermore, toxic levels of copper can be overcome through the *tcr*YAZB gene cluster (27). The co-transfer of copper tolerance with resistance to erythromycin, tetracycline, vancomycin, aminoglycosides or ampicillin resistance has also been demonstrated (28). As such, future studies analysing the antimicrobial efficacy of copper-based surfaces should include the assessment of relevant *Enterococcus* spp.

### *In situ* analysis of coated door handles and panels

Relatively few coated surfaces could be sampled than initially intended due to practicality and logistical issues. The coating process required specialist machinery and took place off-site with a limited throughput. Furthermore, new iterations of coatings were continuously being developed to enhance antimicrobial efficacy, visual quality and mechanical durability. Where the surfaces became aesthetically sub-standard, the door handles had to be replaced.

When evaluating environmental bioburden data, it is important to consider the parameters of cleanliness, a concept of which no definitive definition has been achieved. Early estimates by Griffith, Cooper (29) suggested that based on food preparation standards, an aerobic colony count of greater than 2.5 CFU/cm^2^ should be subject to investigation. Dancer (30) later proposed a 5 CFU/cm^2^ cutoff based on US Department of Agriculture limits of bacteria on food-processing equipment, but has since reverted to 2.5 CFU/cm^2^ in more recent publications (31). A systematic review of copper surfaces by Pineda, Hubbard (32) also identified a threshold of 2.5 CFU/cm^2^ was common practice. However, concerns have been raised as to whether this figure is appropriate, with a large proportion of sites in a dynamic hospital environment likely to fail screening (33). In this study, utilising a 2.5 CFU/cm^2^ cutoff, 45.6% coated and 52.3% uncoated surfaces would have failed screening. Even increasing to 5 CFU/cm^2^ would result in a failure of 33.5% coated and 38.8% uncoated surfaces respectively. A deeper investigation into each of these individual instances of high bioburden would not be feasible.

Overall, median (IQR) counts of 4 (1.2-14.5) CFU/cm^2^ from uncoated and 2 (0.4-7.6) CFU/cm^2^ from coated surfaces highlighted a reduced microbial burden, with our model indicating a 97% probability that uncoated surfaces have higher bacterial loads, albeit with high variability. Whether this provides a tangible reduction in HAIs would require larger scale studies. Our model also indicated a 99% probability of reduced bacterial load on Wednesday. This could be due to the timing of sample collection, as the ward cleaning regimen consisted of daily cleans for high touch surfaces. With a higher number of samples collected on Mondays, sample collection extended later into the day, increasing the opportunity for door use. Further investigation is needed to identify influential factors. This would also clarify the effect of door location, which appeared uncertain. Toilet doors were observed to have above-average CFU counts, although the reasoning behind this was not explored. Nevertheless, this finding highlights a useful, if unsurprising, target for future interventions, including the application of antimicrobial surface coatings and targeted cleaning regimens.

### *In situ* analysis of coated surfaces installed on hospital outpatient check in screens

The mean CFU/cm^2^ recovered from check in screens were consistently lower at all time points than those observed from door handles. Median values displayed the same relationship except for the 09:00 time point. This was hypothesised to be the result of higher traffic in the morning as outpatients checked in for their appointments. However, anecdotal observations suggested that many patients preferred to speak with the receptionist instead of interacting with the screens. We hypothesised that hospital check in screens are not being utilised by many of the patients and hence are not likely to be a high priority in terms of pathogen transmission. It is also plausible that people sterilise their hands with alcohol hand gel upon first entering the hospital prior to interacting with any surface.

## Conclusion

Our study showcased the development of novel antimicrobial surface coatings with a broad spectrum of activity against ESKAPE(e) pathogens. *In situ* analysis of coated door handles identified a large reduction in microbial bioburden, with 77% higher CFU/cm^2^ recovered from uncoated surfaces. The largest CFU counts were observed on toilet door handles, although the extent of this difference relative to other locations was statistically uncertain.

The research and development of novel coatings was ongoing throughout the duration of the project. Due to time and resource limitations, we were unable to assess the coatings over an extended period. To further evidence coating efficacy and economic viability, research over a longer time frame would be required, and correlations with HAIs explored. It would also prove beneficial to identify more evidence-based thresholds for acceptable standards of cleanliness. Previously suggested values were unsuitable for the surfaces assessed within this study.

## Data Availability

All data produced in the present study are available upon reasonable request to the authors.

## Acknowledgements

We would like to thank the Estates teams at the Liverpool School of Tropical Medicine and the Royal Liverpool University Hospital for their support in facilitating the door handle replacements.

## Conflict of interest statement

The authors declare no conflicts of interest.

## Funding Statement

This work was supported by Innovate UK (project no. 10026942) and UKRI through the Strength in Places Fund (grant no. SIPF 36348) under the Infection Innovation Consortium (iiCON) umbrella.

## Notes

### Competing Interest Statement

The authors have declared no competing interest.

