## Supplementary Table A1 for "*In vitro* and *in situ* analysis of a novel copper-based antimicrobial surface coating designed to reduce the microbial bioburden of high touch surfaces in a hospital environment"

(a)

| Model 1 | | | | |
|---|---|---|---|---|
| Parameter | Estimate | 95% CrI | Rhat | Bulk_ESS |
| Intercept | 5.53898537 | [4.28, 6.74] | 1.001902481 | 2816.013034 |
| DayWednesday | -0.591085515 | [-1.13, -0.09] | 1.001278135 | 3501.094698 |
| HandleUncoated | 0.566640803 | [-0.02, 1.16] | 1.001280924 | 3879.405305 |
| Random SD (Date) | 0.610289836 | [0.41, 0.87] | 1.001117852 | 2701.524043 |
| Random SD (Door) | 1.250452846 | [0.63, 2.54] | 1.001158519 | 2884.443245 |
| Shape | 0.382116635 | [0.34, 0.43] | 1.000784947 | 7635.269274 |
| Zero-inflation (zi) | 0.016478764 | [0, 0.04] | 1.000542343 | 5134.180186 |

(b)

| Model 2 | | | | |
|---|---|---|---|---|
| Parameter | Estimate | 95% CrI | Rhat | Bulk_ESS |
| Intercept | 5.525823628 | [5.1, 5.99] | 1.001325227 | 2668.432103 |
| Time11:00 | -0.355319227 | [-0.85, 0.13] | 1.002427904 | 2513.182115 |
| Time13:00 | -1.661875618 | [-2.19, -1.17] | 1.004788374 | 2318.587945 |
| Time15:00 | -0.714385922 | [-1.23, -0.23] | 1.003776999 | 2271.484774 |
| ScreenUncoated | 0.250199058 | [-0.15, 0.64] | 0.999946944 | 4332.320291 |
| Shape | 0.346282921 | [0.28, 0.43] | 1.001528659 | 2113.267739 |
| Zero-inflation (zi) | 0.069401803 | [0.01, 0.13] | 1.003183032 | 1908.07343 |

Supplementary Table A1. Bayesian zero-inflated negative binomial regression model outputs for (a) Model 1 - the fixed effects of the day of sampling (Monday vs Wednesday) and surface type (coated vs uncoated), with random intercepts included for door location and sampling date; and (b) Model 2 - the fixed effects of touchscreen sampling time as a categorical variable (09:00, 11:00, 13:00 and 15:00; 09:00 used as the reference level) and surface type (coated vs uncoated).
