## Supplementary Figure A1 for "*In vitro* and *in situ* analysis of a novel copper-based antimicrobial surface coating designed to reduce the microbial bioburden of high touch surfaces in a hospital environment"

(a)

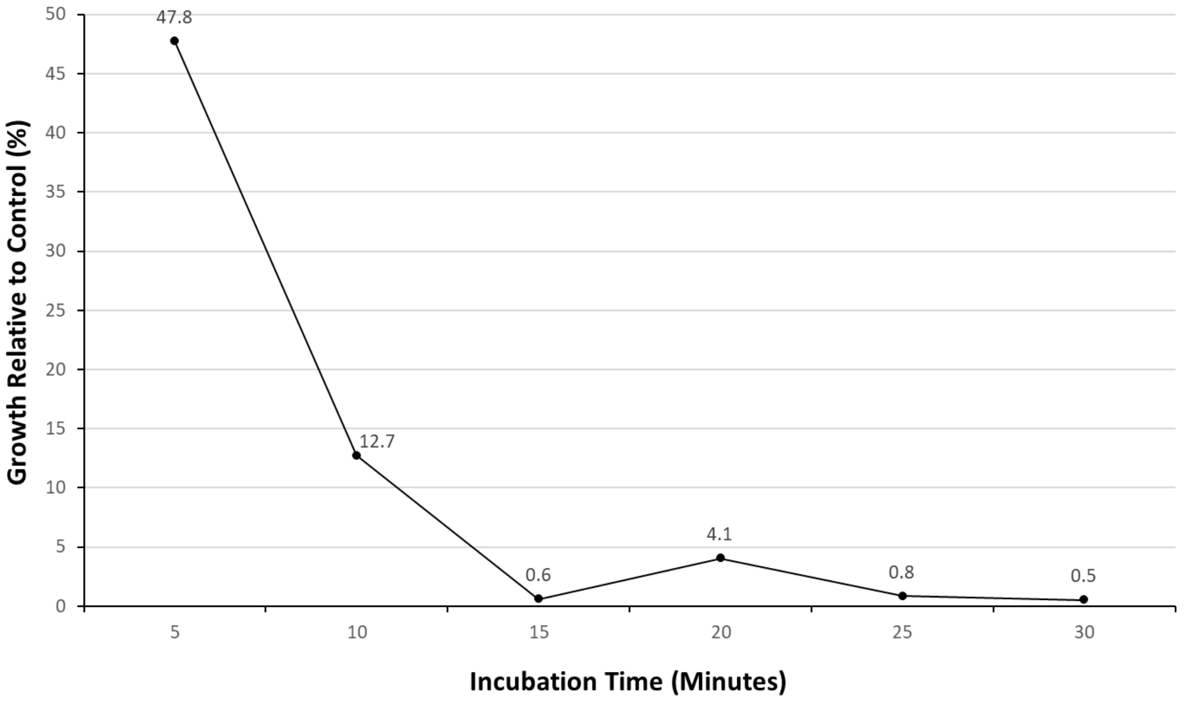


(b)

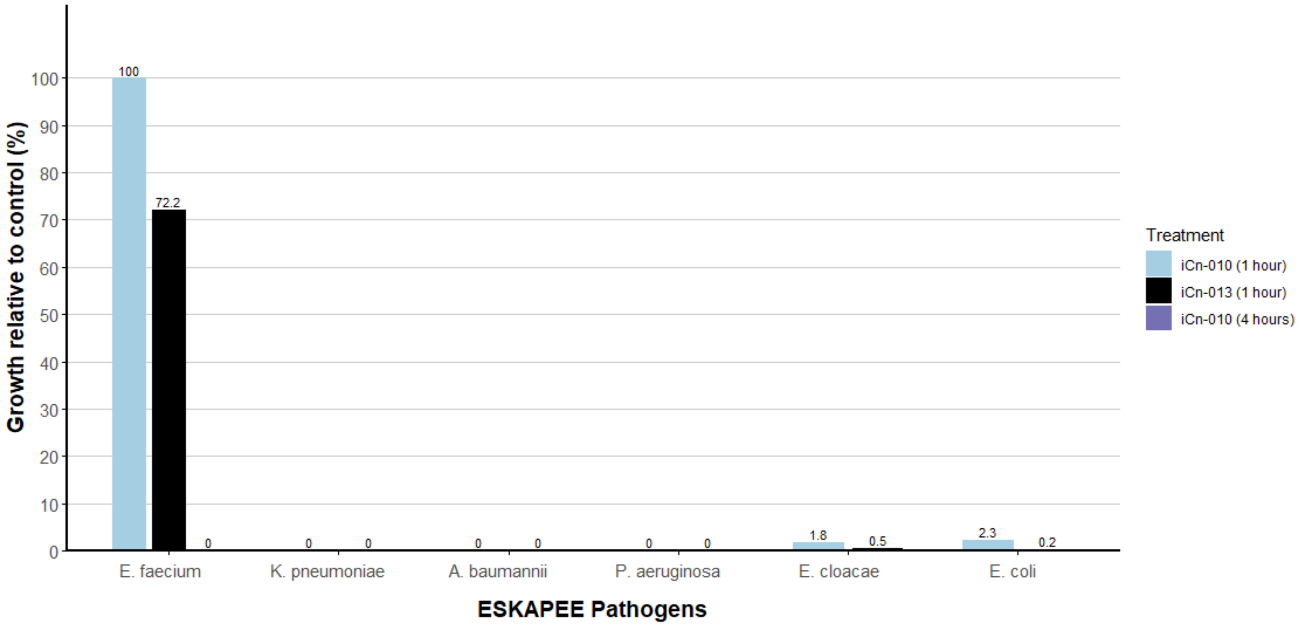


Supplementary Figure A1. (a) The percentage of *Staphylococcus aureus* colony forming units recovered from coating iCn-010 relative to the control after a 5, 10, 15, 20, 25 and 30 minute incubation at room temperature. (b) The percentage of colony forming units recovered from coatings iCn-010 and iCn-013 relative to the control after a 1-hour incubation at room temperature with each of the ESKAPEE pathogens (excluding previously tested *S. aureus*). *Enterococcus faecium* was further tested with a 4-hour incubation period.
